# The relationship between electrophysiological measures of the electrically evoked compound action potential and cochlear implant speech perception outcomes

**DOI:** 10.1101/2022.10.20.22281326

**Authors:** Jeffrey Skidmore, Jacob J. Oleson, Yi Yuan, Shuman He

## Abstract

**Objective:** This study assessed the relationship between electrophysiological measures of the electrically evoked compound action potential (eCAP) and speech perception scores measured in quiet and in noise in post-lingually deafened adult cochlear implant (CI) users. It tested the hypothesis that how well the auditory nerve (AN) responds to electrical stimulation is important for speech perception with a CI in challenging listening conditions.

**Design:** Study participants included 24 post-lingually deafened adult CI users. All participants used Cochlear® Nucleus™ CIs in their test ears. In each participant, eCAPs were measured at multiple electrode locations in response to single-pulse, paired-pulse, and pulse-train stimuli. Independent variables included six parameters calculated from the eCAP recordings: the electrode-neuron interface (ENI) index, the neural adaptation (NA) ratio, NA speed, the adaptation recovery (AR) ratio, AR speed, and the amplitude modulation (AM) ratio. The ENI index quantified the effectiveness of the CI electrodes in stimulating the targeted AN fibers. The NA ratio indicated the amount of NA at the AN caused by a train of constant-amplitude pulses. NA speed was defined as the speed/rate of NA. The AR ratio estimated the amount of recovery from NA at a fixed time point after the cessation of pulse-train stimulation. AR speed referred to the speed of recovery from NA caused by previous pulse-train stimulation. The AM ratio provided a measure of AN sensitivity to AM cues. Participants’ speech perception scores were measured using Consonant-Nucleus-Consonant (CNC) word lists and AzBio sentences presented in quiet, as well as in noise at signal-to-noise ratios (SNRs) of +10 and +5 dB. Predictive models were created for each speech measure to identify eCAP parameters with meaningful predictive power.

**Results:** The ENI index and AR speed had significant bivariate relationships with speech perception scores measured in this study, while the NA ratio, NA speed, the AR ratio, and the AM ratio did not. The ENI index was identified as the only eCAP parameter that had unique predictive power for each of the speech test results. The amount of variance in speech perception scores (both CNC words and AzBio sentences) explained by the eCAP parameters increased with increased difficulty in the listening condition. Over half of the variance in speech perception scores measured in +5 dB SNR noise (both CNC words and AzBio sentences) was explained by a model with only three eCAP parameters: the ENI index, NA speed, and AR speed.

**Conclusions:** The ENI index is the most informative predictor for speech perception performance in CI users. The response characteristics of the AN to electrical stimulation is more important for speech perception with a CI in noise than it is in quiet.

## INTRODUCTION

Over 730,000 individuals who are deaf or severely hard-of-hearing have received a cochlear implant (CI) (National Institute on Deafness and Other Communication Disorders, 2021). Despite general improvements in auditory perception after receiving a CI (e.g., Bittencourt et al., 2012; Boisvert et al., 2020; Hey et al., 2020; Rasmussen et al., 2022), there remains large variability in speech recognition performance among CI patients (e.g., Holden et al., 2013; Blamey et al., 2013; Goudey et al., 2021). While some CI patients can converse without lip-reading, others can only perceive environmental sounds (e.g., Gifford et al., 2008; Holden et al., 2013; Han et al., 2019). Identifying factors accounting for the observed speech perception variability among CI users has been a highly active area of research (e.g., Blamey et al., 1996, 2013; Lazard et al., 2012; Holden et al., 2013, 2016; Kaandorp et al., 2017; James et al., 2019; Zhao et al., 2020; Goudey et al., 2021; Heutink et al., 2021). In general, better speech perception outcomes for CI users have been associated with shorter duration of deafness (Lazard et al., 2012; Blamey et al., 1996, 2013; Zhao et al., 2020; Bernhard et al., 2021; Goudey et al., 2021), better residual hearing before CI (Lazard et al., 2012; Blamey et al., 2013; Boisvert et al., 2020; Zhao et al., 2020; Goudey et al., 2021), closer electrode-to-neuron distance (Finley et al., 2008; Holden et al., 2013, 2016; Heutnik et al., 2021), right-ear implantation (Kraaijenga et al., 2018; Liang et al., 2020; Goudey et al., 2021), and better cognitive function (Holden et al., 2013; Kaandorp et al., 2017; Mussoi & Brown, 2019). However, factors investigated in these studies can only explain approximately 10-40% of the variance in speech perception scores among CI users (Blamey et al., 1996, 2013; Lazard et al., 2012; Holden et al., 2013; James et al., 2019; Zhao et al., 2020; Goudey et al., 2021). Therefore, further studies to identify additional key factors accounting for the speech perception variability among CI users are urgently needed.

CIs provide auditory information to implanted patients by converting acoustic signals into sequences of electrical pulses (i.e., pulse trains) that stimulate nearby auditory nerve (AN) fibers. Subsequently, AN fibers transmit the information to higher-level neural structures for further processing and interpretation. Theoretically, how well electrical stimulation is encoded and processed by the AN should be an important factor for speech perception outcomes in CI users. Results reported in the auditory literature support this theory. Specifically, results of previously published studies suggest that the presence of AN response to electrical stimulation (i.e., the presence of the electrically evoked compound action potential, eCAP), faster recovery from refractoriness, and faster growth of eCAP amplitudes with increasing stimulation level are associated with better speech perception in CI users (for a review, see van Eijl et al., 2017). More recently, it has been shown that better speech perception outcomes are associated with higher effectiveness of the CI electrodes in stimulating the targeted AN fibers (i.e., electrode-to-neuron interface, ENI, Bierer, 2010; Skidmore et al., 2021; Arjmandi et al., 2022), and faster recovery from neural adaptation (NA) induced by prior stimulation (He et al., 2022c). Based on these promising results, we took a bottom-up approach in this study to determine/identify peripheral factors that are important for speech perception outcomes in CI users.

In this study, electrophysiological measures of the eCAP were used to assess the quality of the ENI and several aspects of temporal responsiveness of the AN to electrical stimulation. The eCAP is a neural response that is generated by a population of AN fibers responding synchronously to electrical stimulation (He et al., 2017). Like the ENI, eCAP measures in response to single-pulse and paired-pulse stimulation are affected by electrode position, intracochlear resistance, and the density of AN fibers (e.g., Eisen & Franck, 2004; Shepherd et al., 2004; Brown et al., 2010; Ramekers et al., 2014; Schvartz-Leyzac & Pfingst, 2016; Pfingst et al., 2015, 2017; He et al., 2018; Schvartz-Leyzac et al., 2020). Therefore, eCAPs measured in response to single-pulse and paired-pulse stimulation can be considered as a functional readout for the quality of the ENI (Skidmore et al., 2022a). The temporal response properties of the AN evaluated in this study included NA and recovery from NA (i.e., adaptation recovery, AR) induced by trains of biphasic pulses with constant amplitudes (e.g., Wilson et al., 1997; Hay-McCutcheon et al., 2005; Hughes et al., 2012; Zhang et al., 2013; Ramekers et al., 2015; He et al., 2016; Mussoi & Brown, 2019; He et al., 2022a, b, c, d) and the sensitivity to sinusoidal amplitude modulation (AM) cues implemented in the pulse-train stimulation (Nourski et al., 2007; Tejani et al., 2017; Riggs et al., 2021). NA and AR of the AN were selected as measures of interest because of their essential roles in accurately encoding speech sounds (Delgutte, 1980; Johnson, 1980; Delgutte & Kiang, 1984). AN sensitivity to AM cues was selected as a measure of interest because temporal cues are particularly important for speech perception in CI users and they are encoded in the AM of pulse trains delivered by the CI (Wilson et al., 1991).

The relationship between speech perception outcomes and each of these eCAP parameters has been evaluated in different studies. For example, Skidmore et al. (2021) assessed the correlation between the quality of the ENI, as quantified by the ENI index estimated based on eCAP results, and Consonant-Nucleus-Consonant (CNC) word (Peterson & Lehiste, 1962) and AzBio sentence (Spahr et al., 2012) scores measured in quiet in post-lingually deafened adult CI users. Their results showed a significant, positive correlation between the ENI index and CNC word and AziBio sentence scores. He et al. (2022c) investigated the effects of NA and AR of the AN on CNC word scores measured in quiet and in noise at a signal-to-noise ratio (SNR) of +10 dB in post-lingually deafened adult CI users. Their results showed a nonsignificant relationship between NA of the AN and CNC word scores measured in both conditions. This null finding is consistent with the nonsignificant results reported by Zhang et al. (2013). The speed of AR was found to account for 14.1% of the variability in CNC word scores measured in quiet and 16.7% of the variability in CNC word scores measured in noise with a SNR of +10 dB. This significant finding was not consistent with the results reported by Mussoi & Brown (2019). However, it should be pointed out that AR of the AN was only evaluated at one electrode location for each participant in Mussoi & Brown (2019). Results of a recent study showed that correlating eCAP parameters measured at one electrode location with speech scores can lead to inaccurate conclusions (He et al., 2022d). In addition, the speed of AR was quantified using different methods in these two studies. These methodological differences might account for the inconsistent results reported in Mussoi & Brown (2019) and He et al. (2022c). Finally, He et al. (2022d) evaluated the association between AN sensitivity to AM cues and CNC word scores measured in quiet and in noise at a SNR of +10 dB. Their results revealed a nonsignificant relationship between these measures.

Even though the results of these previous studies are informative and make a scientific contribution to the literature, they have two limitations. First, none of these studies measured all these important eCAP parameters and speech perception scores in the same group of study participants. As a result, no study has included these eCAP parameters in a multiple regression model to predict speech perception scores. Rather, most studies cited above reported bivariate correlations between a single eCAP measure and a speech perception score. These correlation analyses do not account for other eCAP parameters that may explain some of the same variance in speech perception scores. Therefore, the results from correlation analyses may not reflect the unique explanatory power of individual eCAP parameters. Consequently, the results of these studies do not provide conclusive information about which eCAP parameters have the greatest predictive power for speech perception outcomes in CI patients. In addition, speech perception tests implemented in these studies are not representative of current audiological practice for CI patients. Specifically, both CNC word lists and AzBio sentence lists are recommended as speech perception tests for assessing clinical outcomes in adult CI users (American Academy of Audiology, 2019). In addition, assessing speech perception performance in both quiet and in noise testing conditions is recommended (Adunka et al., 2018). The noise conditions with SNRs of +10 and +5 dB are most commonly used in clinical practice for CIs (Carlson et al., 2018). Previous studies either tested speech perception performance only in quiet (Zhang et al., 2013; Skidmore et al., 2021), using another speech test (Mussoi & Brown, 2019), or did not include the noise condition with a SNR of +5 dB (He et al., 2022a, c, d). These caveats limit the clinical application of these previously reported results.

There are some pieces of evidence suggesting that the importance of faithful neural encoding of auditory information at the AN to speech perception may increase as the listening condition becomes more challenging. For example, competing background noise has a much larger, negative effect on speech perception performance in patients with auditory neuropathy spectrum disorder, a disorder characterized by dyssynchrony in AN fiber activity, than in patients with typical sensorineural hearing loss (Starr et al., 1998; Kraus et al., 2000; Shallop, 2002; Zeng & Liu, 2006; Rance et al., 2007; Berlin et al., 2010; Walker et al., 2016). Improving neural synchrony in AN fiber activity using electrical stimulation of a CI could reduce the difference in the magnitude of the noise effect on speech perception performance between these two patient populations (for reviews, see Myers & Nicholson, 2021; Bo et al., 2022). Second, the speed of AR of the AN accounted for more variability in CNC word scores measured at +10 dB SNR than measured in quiet (He et al., 2022c). Unfortunately, these interesting results do not provide conclusive information about the relative importance of AN responsiveness to electrical stimulation to speech perception outcomes in different listening conditions or which AN response properties are more important for listening in challenging conditions than other properties. There is a scientific and clinical need to address these two knowledge gaps.

To address the study limitations and knowledge gaps described above, this study evaluated the association between six eCAP parameters and speech perception scores (CNC words and AzBio sentences) measured in quiet and in noise at SNRs of +10 and +5 dB in post-lingually deafened adult CI users. The primary objective of this study was to identify eCAP parameters that were important predictors for CI speech perception outcomes. We hypothesized that how well the AN encodes and processes electrical stimulation is more important for understanding speech in increased challenging listening conditions. It was expected that the amount of variance in speech perception scores explained by the eCAP parameters (i.e., R^2^s of the predictive models) would be increased as the listening conditions became more challenging due to higher levels of competing background noise.

## MATERIALS AND METHODS

### Study Participants

Study participants included 24 post-lingually deafened adult CI users. All participants were implanted with a Cochlear™ Nucleus® device (Cochlear Ltd., Sydney, NSW, Australia) with a full electrode insertion. Only one ear was tested for each participant. For bilateral CI users, the test ear was selected pseudo-randomly. Written informed consent was obtained from all study participants at the time of data collection. The study was approved by the Biomedical Institutional Review Board (IRB) at The Ohio State University (IRB study #: 2017H0131).

### eCAP Measurements and Parameters

The procedures for obtaining the eCAP were the same as those used in our recent studies (Skidmore et al., 2021; Riggs et al., 2021; He et al., 2022a, b). All eCAPs were obtained using the Advanced Neural Response Telemetry function via the Custom Sound EP (v. 5.1 or 6.0) software interface (Cochlear Ltd, Sydney, NSW, Australia). The stimulus consisted of one or more symmetric, cathodic-leading, biphasic pulses with an interphase gap of 7 µs and a pulse phase duration of 25 µs/phase. For measuring NA and AR of the AN, the stimulus was a 100-ms pulse train with a pulse rate of 900 pulses per second (pps) per channel. For measuring AN sensitivity to AM cues, the stimulus was a 200-ms pulse train with a carrier rate of 2000 pps that was sinusoidally amplitude modulated at 20 Hz with a modulation depth of 100%. All stimuli were presented in a monopolar-coupled stimulation mode to individual CI electrodes via an N6 sound processor connected to a programming pod.

#### The ENI index

The ENI index is a number between 0 and 100 that represents the overall quality of the ENI, where larger numbers represent better ENIs. The ENI index was calculated for each participant using the model created by Skidmore et al. (2021). Briefly, the ENI index was a compound metric composed of four parameters derived from eCAPs evoked by single-pulse and paired-pulse stimulation. The four parameters included the lowest stimulation level that evoked an eCAP (i.e., the eCAP threshold), the slope of the eCAP amplitude growth function, the latency of the first negative peak (i.e., N1 latency) in the recorded waveform with the largest eCAP amplitude, and the absolute refractory period estimated from the eCAP refractory recovery function. Each of the four parameters was recorded at three electrode locations along the electrode array. These twelve measures (4 parameters x 3 electrode locations) were inputs into the model that was created using linear regression with elastic net regularization to produce the ENI index. Additional details regarding the ENI index (originally called the cochlear nerve index), including a detailed description of its development, are contained in Skidmore et al. (2021).

#### Neural adaptation of the AN

Details of using eCAPs measures to assess NA of the AN have been reported in He et al. (2022a). Briefly, eCAPs evoked by pulses 1-8, 40-45, and 85-89 in a 100-ms pulse-train stimulus were recorded. Smaller eCAP amplitudes were evoked by the pulses toward the end of the pulse train due to the AN fibers gradually adapting to the constant-amplitude stimulus.

##### The NA ratio

The NA ratio is a measure of the amount of NA that occurs in response to a train of constant amplitude pulses, with smaller NA ratios indicating greater NA. The NA ratio was calculated at individual electrode locations by averaging normalized amplitudes (re: the eCAP amplitude evoked by a single pulse stimulus presented at the same stimulation level) of eCAPs evoked by pulses 1-8, 40-45, and 85-89. The NA ratio was calculated based on eCAP results measured at four electrode locations along the electrode array, and then averaged together to create one measure for each participant in this study.

##### NA speed

NA speed was estimated using a two-parameter power law function in the form of

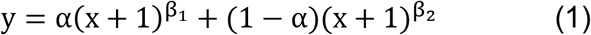

where y represented the normalized eCAP amplitude (re: the eCAP amplitude evoked by a single pulse) evoked by the last pulse of a pulse-train of duration x. β_1_ and β_2_ represented the time constant of NA occurring within the first 8-ms of stimulation (i.e., short phase) and between 8 and 100 ms of stimulation (i.e., long phase), respectively. α indicated the relative contribution of the short phase and the long phase to the overall fit of the function. For this study, NA speed was calculated by averaging the absolute values of β_1_ estimated at four electrode locations along the electrode array. Larger values of the averaged absolute values of β_1_ indicated faster NA.

#### Neural Adaptation Recovery of the AN

Details of using eCAPs measures to assess the amount and the speed of AR are reported in He et al. (2022b). Briefly, eCAPs evoked by the probe pulse presented at 1.054, 2, 4, 8, 16, 32, 64, 128 and 256 ms after the offset of the last pulse of the masker pulse-train were recorded. As the masker-probe-interval (MPI) increased, AN fibers gradually recovered from the NA induced by the pulse-train masker, which resulted in gradually increased eCAP amplitudes at longer MPIs.

##### The AR ratio

The AR ratio quantifies the amount of recovery from NA induced by a constant-amplitude pulse-train. Larger AR ratios indicate greater recovery from NA. The AR ratio was calculated by dividing the amplitude of the eCAP to the probe pulse presented after 256 ms of the end of the pulse-train stimulation (i.e., MPI = 256 ms) by the eCAP amplitude evoked by a single-pulse stimulus presented at the same stimulation level. In this study, the AR ratio was calculated at four electrode locations along the electrode array, and then averaged together to create one measure for each participant.

##### AR speed

AR speed was estimated using a mathematical model with up to three exponential components in the form of

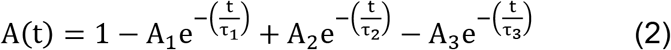

where A(t) represented the normalized eCAP amplitude (re: the eCAP amplitude evoked by a single pulse) evoked by the probe pulse at an MPI of t. A_1_, A_2_ and A_3_ were fitting coefficients and τ_1_, τ_2_ and τ_3_ were the time constants for early enhancement, slow decay, and adaptation recovery, respectively. Detailed descriptions and explanations of these three phases have been reported in He et al. (2022b). For this study, AR speed was calculated by averaging the values of τ_3_ estimated at four electrode locations along the electrode array. Larger averaged τ_3_s indicated slower recovery from NA.

#### The AM ratio

The AM ratio is a measure of AN sensitivity to AM cues, with larger AM ratios indicating greater AN sensitivity to AM cues. The AM ratio was calculated using the same procedures as detailed in Riggs et al. (2021). Briefly, eCAPs evoked by twenty pulses within the last two cycles of an AM pulse train were measured. eCAPs evoked by single pulses with probe levels corresponding to the stimulation levels of the twenty pulses of the AM pulse train were also measured. The AM ratio was then calculated at individual electrode locations as the ratio of the difference in the maximum and the minimum eCAP amplitude measured for the AM pulse train stimulus and the difference in the maximum and the minimum eCAP amplitude measured for the single pulse stimulus. The AM ratio was calculated at seven electrode locations along the electrode array, and then averaged together to create one measure for each participant in this study.

### Speech Measures

Speech perception performance was evaluated using CNC word (Peterson & Lehiste, 1962) and AzBio sentence (Spahr et al., 2012) lists presented in quiet and in two noise conditions. All auditory stimuli were presented in a sound-proof booth via a speaker placed one meter in front of the participant at zero degrees azimuth. The target stimulus was always presented at 60 dB(A) sound pressure level (SPL). For the noise conditions, speech-shaped noise was presented concurrently with the target stimulus at 50 dB(A) SPL or 55 dB(A) SPL (i.e., SNR of +10 dB or +5 dB, respectively).

### Statistical Analyses

For each speech perception test result measured in each listening condition, three types of predictive models were created. First, six individual models were created by using simple linear regression with each eCAP parameter as the only predictor to identify the bivariate relationships between each eCAP parameter and each speech measure. The R^2^ for each individual model indicated the variance in speech scores explained by that parameter without adjusting for the other eCAP parameters. Second, a complete model was created using multiple linear regression with all six eCAP parameters as predictors. The R^2^ for the complete model quantified the contribution of AN responsiveness to speech perception scores by indicating the variance in speech scores explained by all six eCAP parameters together. Third, a reduced model was created by including only the eCAP parameters that explained sufficient and unique variance in speech perception scores as predictors in the multiple linear regression analysis. All possible combinations of the eCAP parameters were evaluated and the combination of eCAP parameters with the lowest Akaike Information Criterion (Akaike, 1974) was selected as the reduced model. The R^2^ for the reduced model indicated the variance in speech scores explained by the eCAP parameters selected in the model. The assumptions of regression analyses were evaluated by examining the residuals of all models created in this study and no violations were detected. All statistical modeling was performed using R v. 4.2 (R Core Team, 2022) with a 0.05 level of significance.

## RESULTS

Results of CNC word tests and AzBio sentence tests as a function of six eCAP parameters for each of the three listening conditions are shown in Figure 1 and Figure 2, respectively. Overall, speech perception performance decreased with increasing level of competing background noise. Additionally, visual inspection of these figures revealed substantial variations in the bivariate relationship between each eCAP parameter and each speech test result.

**Figure 1.**
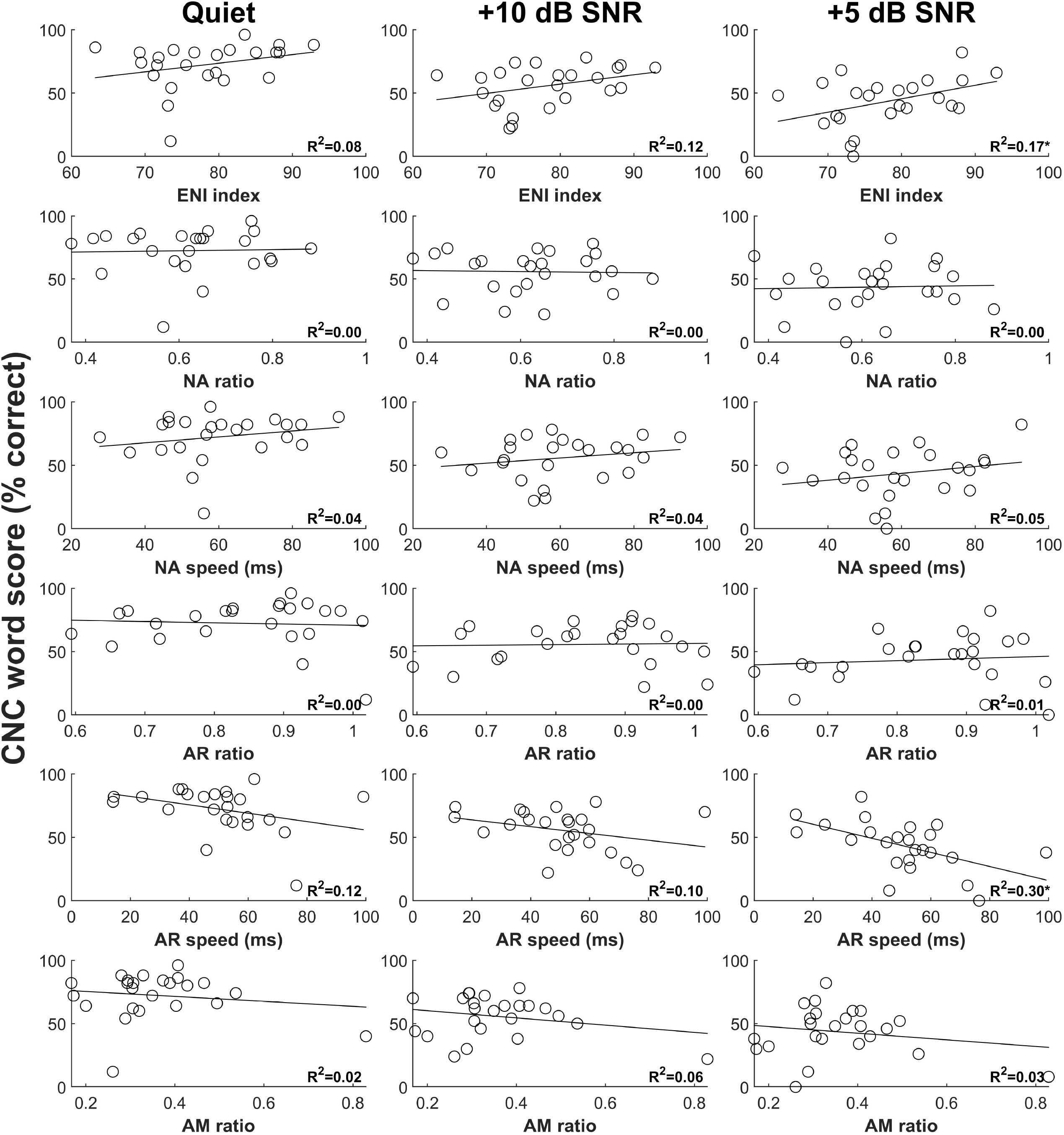
Results of CNC word tests as a function of six eCAP parameters for three listening conditions. Each circle represents the result for an individual study participant. The variance in CNC word scores explained by the eCAP parameter calculated from simple linear regression is provided in the lower right-hand corner of each panel. Statistically significant results from the regression analyses are indicated by an asterisk. CNC, Consonant-Nucleus-Consonant; eCAP, electrically evoked compound action potential.

**Figure 2.**
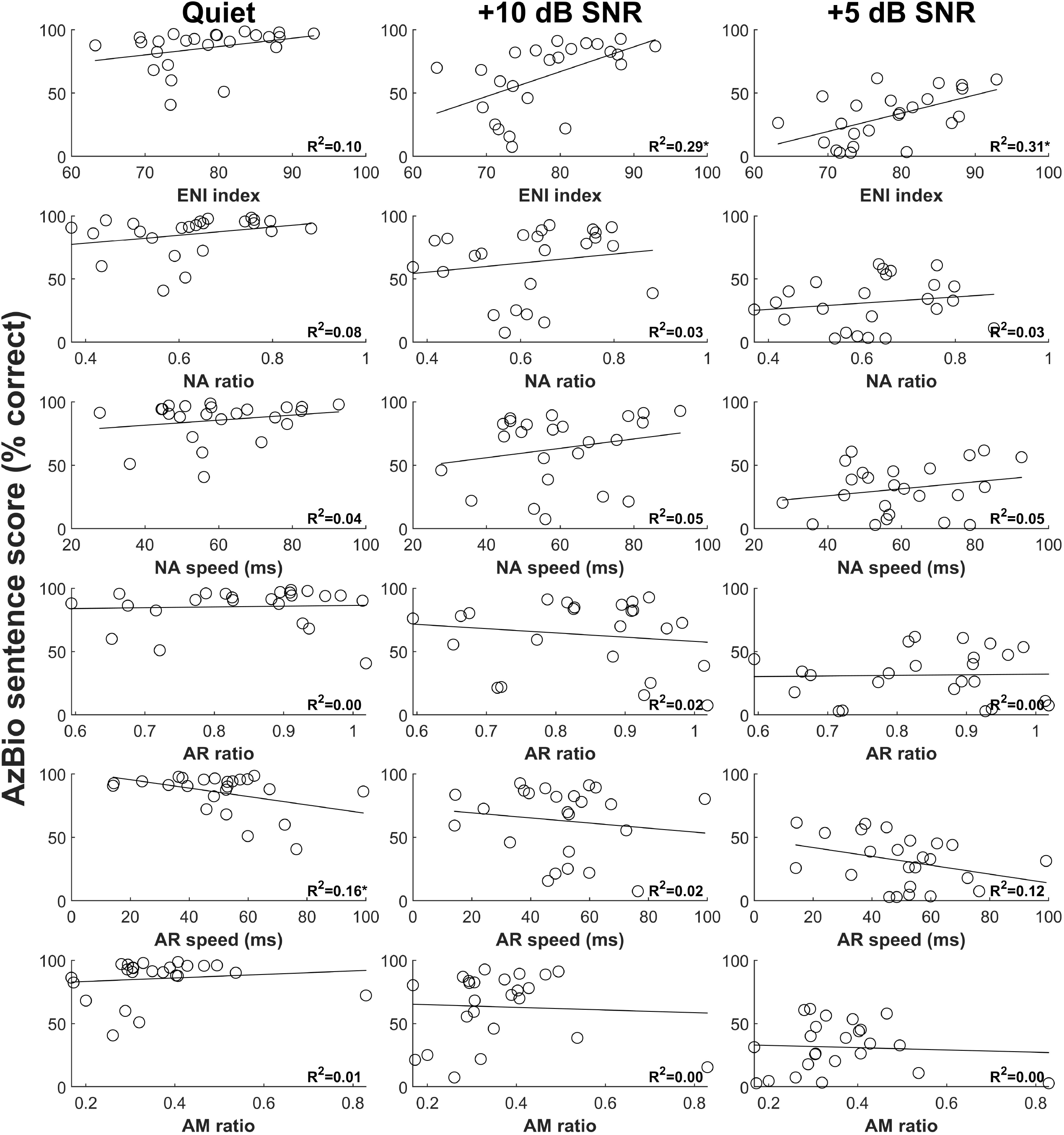
Results of AzBio sentence tests as a function of six eCAP parameters for three listening conditions. Each circle represents the result for an individual study participant. The variance in AzBio sentence scores explained by the eCAP parameter calculated from simple linear regression is provided in the lower right-hand corner of each panel. Statistically significant results from the regression analyses are indicated by an asterisk. eCAP, electrically evoked compound action potential.

### Individual Models

The results of each individual model assessing the bivariate relationship between an individual eCAP parameter and a speech test result clearly indicated that there were no significant relationships between any of the eCAP parameters and CNC word scores measured in quiet or in +10 dB SNR noise (see Table A1). However, the ENI index and AR speed were significantly related with CNC word scores measured in +5 dB SNR noise (ENI index: β = 1.06, t = 2.12, p = 0.046, R^2^ = 0.17); AR speed: β = -0.56, t = -3.06, p = 0.006, R^2^ = 0.30). For AzBio sentences (see Table A2), AR speed was the only eCAP parameter that was significantly related with scores measured in quiet (β = -0.33, t = - 2.08, p = 0.049, R^2^ = 0.16), and the ENI index was the only eCAP parameter significantly related with scores measured in both noise conditions (+10 dB SNR: β = 1.96, t = 2.98, p = 0.007, R^2^ = 0.29; +5 dB SNR: β = 1.44, t = 3.11, p = 0.005, R^2^ = 0.31). For all significant relationships, better speech performance was associated with better quality of the ENI (i.e., larger ENI indices) and faster AR (i.e., smaller values of AR speed).

### Complete Models

The results of the complete models revealed that the variance in scores on each speech test explained by the eCAP parameters increased with increased difficulty in the listening condition (see Table B1 and Table B2). In the most difficult listening condition (i.e., +5 dB SNR), the eCAP parameters explained over half of the variance in both CNC word scores (R^2^ = 0.57) and AzBio sentence scores (R^2^ = 0.51).

### Reduced Models

The results of each reduced model that only included the eCAP parameters that contributed unique power to predicting CNC word scores and AzBio sentence scores are provided in Table 1 and Table 2, respectively. As shown in the tables, the ENI index was the only eCAP parameter that was selected in each of the six reduced models. Therefore, the ENI index contributed unique predictive power for each of the six speech test results that was independent of the other eCAP parameters. Along with the ENI index, NA speed and AR speed were selected in both reduced models for speech scores measured in +5 dB SNR noise. In this noise condition, these three eCAP parameters explained 53% and 51% of the variance in CNC word scores and AzBio sentence scores, respectively. Also, the R^2^ values of the reduced models increased with increased difficulty in the listening condition.

**TABLE 1.** Results of statistical models that only included eCAP parameters that contributed unique predictive power to explaining variance in CNC word scores measured in three listening conditions. eCAP, electrically evoked compound action potential; CNC, Consonant-Nucleus-Consonant; SNR, signal-to-noise ratio; ENI, electrode neuron interface; NA, neural adaptation; AR, adaptation recovery; AM, amplitude modulation.

| Listening condition | eCAP parameter | $\beta$ value | T value | P value | R<sup>2</sup> |
| --- | --- | --- | --- | --- | --- |
| Quiet | ENI index | 0.68 | 1.45 | 0.161 | 0.20 |
|  | AR speed | -0.33 | -1.79 | 0.087 |  |
| +10 dB SNR | ENI index | 0.68 | 1.67 | 0.111 | 0.29 |
|  | AR speed | -0.30 | -1.87 | 0.077 |  |
|  | AM ratio | -30.21 | -1.35 | 0.193 |  |
| +5 dB SNR | ENI index | 1.18 | 2.92 | 0.008 | 0.53 |
|  | NA speed | 0.30 | 1.62 | 0.122 |  |
|  | AR speed | -0.53 | -3.41 | 0.003 |  |

**TABLE 2.** Results of statistical models that only included eCAP parameters that contributed unique predictive power to explaining variance in AzBio sentence scores measured in three listening conditions. eCAP, electrically evoked compound action potential; SNR, signal-to-noise ratio; ENI, electrode neuron interface; NA, neural adaptation; AR, adaptation recovery; AM, amplitude modulation.

| Listening condition | eCAP parameter | $\beta$ value | T value | P value | $R^2$ |
| --- | --- | --- | --- | --- | --- |
| Quiet | ENI index | 0.66 | 1.69 | 0.105 | 0.26 |
|  | AR speed | -0.33 | -2.17 | 0.042 |  |
| +10 dB SNR | ENI index | 2.15 | 3.39 | 0.003 | 0.38 |
|  | NA speed | 0.53 | 1.81 | 0.084 |  |
| +5 dB SNR | ENI index | 1.57 | 3.79 | 0.001 | 0.51 |
|  | NA speed | 0.36 | 1.86 | 0.078 |  |
|  | AR speed | -0.32 | -2.01 | 0.059 |  |

### Summary of Study Results

The ENI index and AR speed were the only two eCAP parameters that had statistically significant bivariate relationships with speech perception scores measured in this study. The amount of variance in speech scores (both CNC words and AzBio sentences) explained by the eCAP parameters increased with increased levels of competing background noise. The ENI index was the only eCAP parameter that was selected in each of the six reduced models. Over half of the variance in speech scores (both CNC words and AzBio sentences) measured in +5 dB SNR noise was explained by only three eCAP parameters: the ENI index, NA speed, and AR speed. These main findings from all the statistical models created in this study are provided in Table 3.

**TABLE 3.**
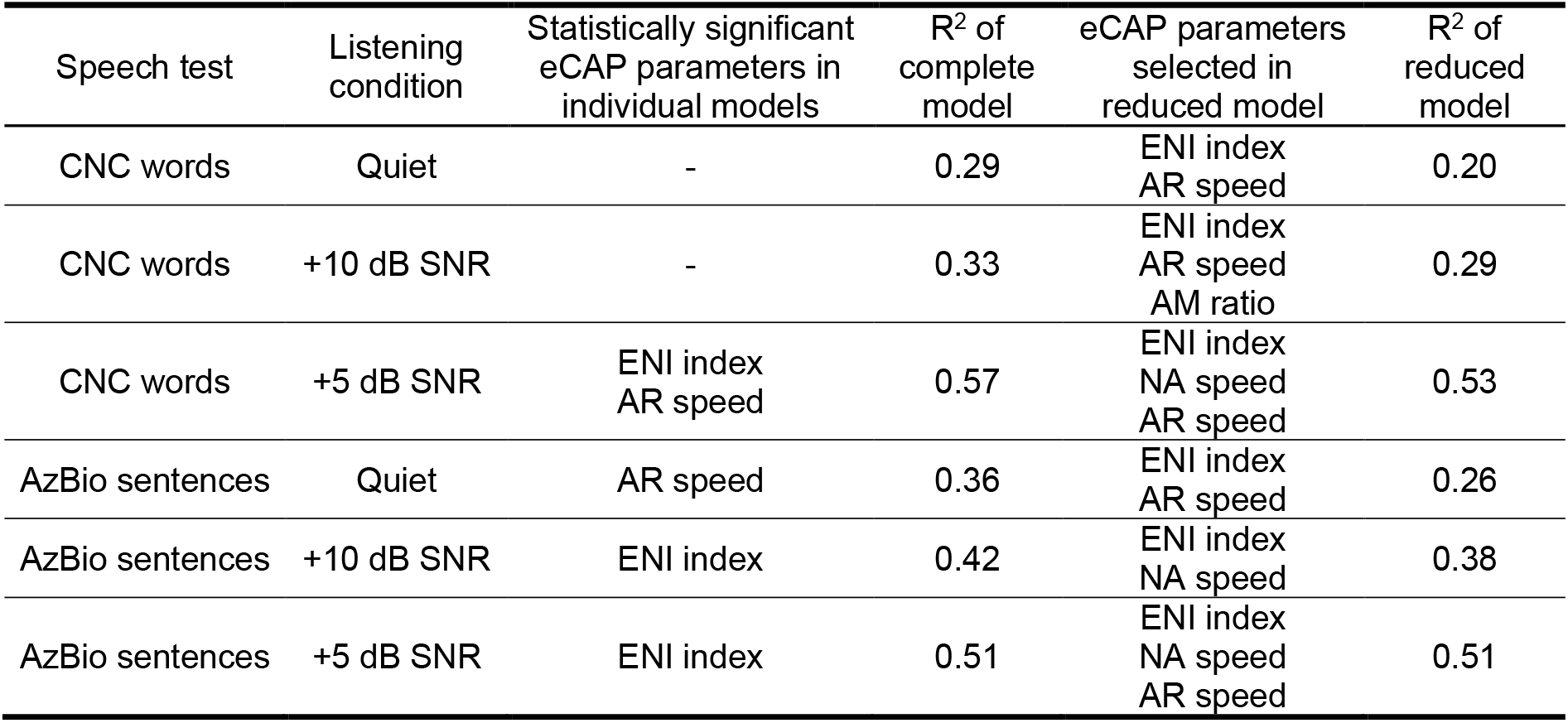
Summary of results from all statistical models in this study. eCAP, electrically evoked compound action potential; CNC, Consonant-Nucleus-Consonant; SNR, signal-to-noise ratio; ENI, electrode neuron interface; NA, neural adaptation; AR, adaptation recovery; AM, amplitude modulation.

| Speech test | Listening condition | Statistically significant eCAP parameters in individual models | R<sup>2</sup> of complete model | eCAP parameters selected in reduced model | R<sup>2</sup> of reduced model |
| --- | --- | --- | --- | --- | --- |
| CNC words | Quiet | - | 0.29 | ENI index<br>AR speed | 0.20 |
| CNC words | +10 dB SNR | - | 0.33 | ENI index<br>AR speed<br>AM ratio | 0.29 |
| CNC words | +5 dB SNR | ENI index<br>AR speed | 0.57 | ENI index<br>NA speed<br>AR speed | 0.53 |
| AzBio sentences | Quiet | AR speed | 0.36 | ENI index<br>AR speed | 0.26 |
| AzBio sentences | +10 dB SNR | ENI index | 0.42 | ENI index<br>NA speed | 0.38 |
| AzBio sentences | +5 dB SNR | ENI index | 0.51 | ENI index<br>NA speed<br>AR speed | 0.51 |

## DISCUSSION

### Comparison of eCAP Parameters

The primary objective of this study was to identify eCAP parameters that were important predictors for speech perception scores in post-lingually deafened adult CI users. Overall, the results of this study showed that the ENI index was the most sensitive predictor of speech perception scores in this patient population. This was most clearly shown by the result of the ENI index being the only eCAP parameter selected in all reduced models. This suggested that the ENI index contributed explanatory power to the predictive capability of all the models that was independent of the other eCAP parameters. Additionally, the ENI index had a significant bivariate relationship with three of the speech test results, which was more than any other eCAP parameter. This result demonstrating the value of the ENI index in predicting speech perception scores is generally consistent with what has been reported in the literature. Specifically, better speech perception outcomes have been reported in CI users with higher quality ENI as estimated by electrode placement (Finley et al., 2008; Heutink et al., 2021), size of the AN in imaging results (Kang et al., 2010), psychophysical detection thresholds (Garadat et al., 2013), focused stimulation thresholds (Long et al., 2014), and eCAP measures (Skidmore et al., 2021).

While not the most sensitive predictor of speech scores in this study, AR speed also had meaningful predictive power in the models. This was shown by AR speed being selected in five of the reduced models and having a significant bivariate relationship with two of the speech test results. This result is also supported by our recent study in which we reported a moderate, negative correlation between CNC word scores measured in quiet and in noise and the speed of AR averaged across multiple electrodes for each participant (He et al., 2022c).

In contrast to the ENI index and AR speed, the other eCAP parameters (i.e., the NA ratio, NA speed, the AR ratio and the AM ratio) did not have significant relationships with any of the speech perception scores measured in this study. These results are consistent with the null results from other studies (Zhang et al., 2013; He et al., 2022c, d). Additionally, the NA ratio and the AR ratio were not selected in any of the reduced models, and the AM ratio was selected in only one reduced model, which suggested that these eCAP measures did not contribute a meaningful amount of unique information to the predictive models. However, NA speed was selected in the reduced model for CNC words measured in +5 dB SNR noise and for AzBio sentences tested at SNRs of +10 and +5 dB. Therefore, a measure of the speed of NA may provide beneficial, predictive information for speech perception scores measured in challenging listening conditions, even if it is not a robust predictor of speech perception scores as an individual predictor. This is supported by the idea that normal NA of the AN increases the spectral contrast between successive speech segments and improves the temporal precision of speech onset cues (Delgutte, 1997), which are particularly important for speech perception in challenging listening conditions.

### Comparison of Listening Conditions

This study tested the hypothesis that AN responsiveness to electrical stimulation is especially important for speech perception in challenging listening conditions. The results of this study are consistent with this hypothesis. Specifically, the variances in speech perception scores explained by the eCAP parameters all together (i.e., R^2^s of the complete models) were higher for speech perception scores measured in noise than in quiet. Additionally, the R^2^s of the complete models increased with increased noise for both word and sentence scores. These results suggest that AN responsiveness to electrical stimulation is more important for speech perception in noise compared to speech perception in quiet. We postulate that this phenomenon occurs because less information from the peripheral auditory system may be needed to understand speech in quiet than in noise. Consequently, degraded auditory input to the central auditory system resulting from impaired peripheral encoding would have less detrimental effects on speech understanding in quiet. However, this postulation has not been verified and therefore remains as speculation. A future study will evaluate the relative contribution of peripheral and central factors to speech perception with a CI in quiet and in noise.

### Clinical Application and Implication

Results of this study showed that the ENI index was the most informative predictor for speech perception performance in post-lingually deafened adult CI users. This finding generally aligns with the positive relationship between the quality of ENI and speech perception outcomes reported in CI patients (Finley et al., 2008; Kang et al., 2010; Garadat et al., 2013; Long et al., 2014; Heutink et al., 2021; Skidmore et al., 2021). As a result, the ENI index can potentially be used as a biomarker for predicting CI clinical outcomes.

Results of this study also suggest the importance of enhancing the quality of the ENI for improving speech perception outcomes in CI patients. The quality of the ENI is negatively impacted by poor AN function (Skidmore et al., 2021), bone and tissue growth caused by intracochlear surgical trauma (Seyyedi & Nadol, 2014; Kamakura & Nadol, 2016), and large distances between CI electrodes and their target AN fibers (Finley et al., 2008; Heutink et al., 2021). Therefore, technologies/strategies that better preserve the functional integrity of the AN, reduce surgical trauma and/or improve placement of the electrode array in the cochlea should lead to enhanced ENI quality, and, therefore, result in improved CI outcomes. Dexamethasone-eluting electrode arrays and using robotics-assistance to provide a slow, steady electrode insertion during CI surgery are two novel technologies/strategies along this line that can potentially result in improved CI outcomes.

### Study Limitations

One potential limitation of the present study is that only 24 post-lingually deafened adult participants were included in the study. Therefore, the variance in speech perception performance explained by the eCAP parameters in this study cannot be assumed to represent the variance explained in the entire CI patient population. However, the purpose of this study was to identify the most relevant predictors of speech outcomes for adult CI users, which was accomplished in this study. A future study will evaluate the variance in speech perception scores accounted for by the ENI index and AR speed in a large sample of patients to obtain a more representative estimate of the variance in speech perception scores explained by these two eCAP parameters.

The other potential limitation of the present study is that exclusively eCAP measures were included in the predictive models. Other factors, such as cognition, etiology of hearing loss, and duration of deafness, have been shown to be correlated with speech perception outcomes in CI patients (e.g., Lazard et al., 2012; Blamey et al., 2013; Holden et al., 2013; Kaandorp et al., 2017; Mussoi & Brown, 2019; Zhao et al., 2020; Bernhard et al., 2021; Goudey et al., 2021). Therefore, including these factors should improve the ability of a model to predict speech perception scores. However, the objective of this study was not to create a model that could explain as much variance in speech perception scores as possible. Rather, the objective was to identify the most relevant eCAP predictors of speech. These results provided a foundation for future studies that combine a small subset of eCAP measures (e.g., ENI index and AR speed) with other factors (e.g., cortical encoding and processing of electrical stimulation, cognitive measures, etc.) to better understand the contribution of each level of the neural system to speech perception outcomes in CI patients.

## CONCLUSIONS

The quality of the ENI is the most sensitive predictor of speech perception scores in post-lingually deafened adult CI users, followed by the speed of recovery from NA. A predictive model with three eCAP parameters can explain approximately half of the variance in speech perception scores measured in noise. The responsiveness of the AN to electrical stimulation considerably impacts speech perception outcomes with a CI, especially in difficult listening conditions.

## Data Availability

Please contact the corresponding author to discuss access to the data presented in this study.

## FIGURE LEGENDS

**TABLE A1.** Results of statistical models assessing the bivariate relationships between eCAP parameters and CNC word scores measured in three listening conditions. Statistically significant results are indicated with bold text. eCAP, electrically evoked compound action potential; CNC, Consonant-Nucleus-Consonant; SNR, signal-to-noise ratio; ENI, electrode neuron interface; NA, neural adaptation; AR, adaptation recovery; AM, amplitude modulation.

| Listening condition | eCAP parameter | $\beta$ value | T value | P value | R<sup>2</sup> |
| --- | --- | --- | --- | --- | --- |
| Quiet | ENI index | 0.68 | 1.38 | 0.182 | 0.08 |
|  | NA ratio | 4.67 | 0.16 | 0.872 | 0.00 |
|  | NA speed | 0.23 | 0.99 | 0.331 | 0.04 |
|  | AR ratio | -9.83 | -0.31 | 0.760 | 0.00 |
|  | AR speed | -0.33 | -1.75 | 0.095 | 0.12 |
|  | AM ratio | -19.57 | -0.70 | 0.490 | 0.02 |
| +10 dB SNR | ENI index | 0.73 | 1.72 | 0.100 | 0.12 |
|  | NA ratio | -3.79 | -0.15 | 0.883 | 0.00 |
|  | NA speed | 0.21 | 0.99 | 0.332 | 0.04 |
|  | AR ratio | 4.93 | 0.18 | 0.863 | 0.00 |
|  | AR speed | -0.27 | -1.58 | 0.127 | 0.10 |
|  | AM ratio | -19.57 | -0.70 | 0.490 | 0.02 |
| +5 dB SNR | **ENI index** | **1.06** | **2.12** | **0.046** | **0.17** |
|  | NA ratio | 5.07 | 0.16 | 0.871 | 0.00 |
|  | NA speed | 0.27 | 1.08 | 0.290 | 0.05 |
|  | AR ratio | 16.04 | 0.47 | 0.642 | 0.01 |
|  | **AR speed** | **-0.56** | **-3.06** | **0.006** | **0.30** |
|  | AM ratio | -26.14 | -0.88 | 0.388 | 0.03 |

**TABLE A2.** Results of statistical models assessing the bivariate relationships between eCAP parameters and AzBio sentence scores measured in three listening conditions. Statistically significant results are indicated with bold text. eCAP, electrically evoked compound action potential; SNR, signal-to-noise ratio; ENI, electrode neuron interface; NA, neural adaptation; AR, adaptation recovery; AM, amplitude modulation.

| Listening condition | eCAP parameter | $\beta$ value | T value | P value | R<sup>2</sup> |
| --- | --- | --- | --- | --- | --- |
| Quiet | ENI index | 0.66 | 1.56 | 0.133 | 0.10 |
|  | NA ratio | 32.77 | 1.37 | 0.183 | 0.08 |
|  | NA speed | 0.20 | 0.99 | 0.332 | 0.04 |
|  | AR ratio | 6.22 | 0.23 | 0.823 | 0.00 |
|  | **AR speed** | **-0.33** | **-2.08** | **0.049** | **0.16** |
|  | AM ratio | 13.83 | 0.57 | 0.572 | 0.01 |
| +10 dB SNR | **ENI index** | **1.96** | **2.98** | **0.007** | **0.29** |
|  | NA ratio | 35.76 | 0.83 | 0.413 | 0.03 |
|  | NA speed | 0.37 | 1.05 | 0.306 | 0.05 |
|  | AR ratio | -33.63 | -0.71 | 0.488 | 0.02 |
|  | AR speed | -0.20 | -0.67 | 0.512 | 0.02 |
|  | AM ratio | -10.49 | -0.25 | 0.807 | 0.00 |
| +5 dB SNR | **ENI index** | **1.44** | **3.11** | **0.005** | **0.31** |
|  | NA ratio | 24.52 | 0.80 | 0.433 | 0.03 |
|  | NA speed | 0.28 | 1.09 | 0.287 | 0.05 |
|  | AR ratio | 4.67 | 0.14 | 0.894 | 0.00 |
|  | AR speed | -0.35 | -1.70 | 0.103 | 0.12 |
|  | AM ratio | -9.04 | -0.30 | 0.769 | 0.00 |

**TABLE B1.** Results of statistical models assessing the variance in CNC word scores measured in three listening conditions that was explained by six eCAP parameters. eCAP, electrically evoked compound action potential; CNC, Consonant-Nucleus-Consonant; SNR, signal-to-noise ratio; ENI, electrode neuron interface; NA, neural adaptation; AR, adaptation recovery; AM, amplitude modulation.

| Listening condition | eCAP parameter | $\beta$ value | T value | P value | R<sup>2</sup> |
| --- | --- | --- | --- | --- | --- |
| Quiet |  |  |  |  | 0.29 |
|  | ENI index | 0.67 | 1.19 | .252 |  |
|  | NA ratio | 4.99 | 0.14 | .888 |  |
|  | NA speed | 0.24 | 1.00 | .330 |  |
|  | AR ratio | -20.04 | -0.61 | .551 |  |
|  | AR speed | -0.37 | -1.79 | .091 |  |
|  | AM ratio | -19.56 | -0.60 | .554 |  |
| +10 dB SNR |  |  |  |  | 0.33 |
|  | ENI index | 0.81 | 1.68 | 0.111 |  |
|  | NA ratio | -6.93 | -0.23 | 0.819 |  |
|  | NA speed | 0.22 | 1.08 | 0.294 |  |
|  | AR ratio | 3.31 | 0.12 | 0.908 |  |
|  | AR speed | -0.27 | -1.54 | 0.141 |  |
|  | AM ratio | -24.53 | -0.88 | 0.389 |  |
| +5 dB SNR |  |  |  |  | 0.57 |
|  | ENI index | 1.15 | 2.43 | 0.026 |  |
|  | NA ratio | -4.29 | -0.15 | 0.885 |  |
|  | NA speed | 0.28 | 1.42 | 0.173 |  |
|  | AR ratio | 2.84 | 0.10 | 0.919 |  |
|  | AR speed | -0.56 | -3.24 | 0.005 |  |
|  | AM ratio | -26.16 | -0.97 | 0.348 |  |

**TABLE B2.** Results of statistical models assessing the variance in AzBio sentence scores measured in three listening conditions that was explained by six eCAP parameters. eCAP, electrically evoked compound action potential; SNR, signal-to-noise ratio; ENI, electrode neuron interface; NA, neural adaptation; AR, adaptation recovery; AM, amplitude modulation.

| Listening condition | eCAP parameter | $\beta$ value | T value | P value | $R^2$ |
| --- | --- | --- | --- | --- | --- |
| Quiet |  |  |  |  | 0.36 |
|  | ENI index | 0.61 | 1.31 | 0.206 |  |
|  | NA ratio | 21.28 | 0.74 | 0.468 |  |
|  | NA speed | 0.23 | 1.20 | 0.247 |  |
|  | AR ratio | -10.43 | -0.39 | 0.705 |  |
|  | AR speed | -0.32 | -1.91 | 0.073 |  |
|  | AM ratio | 5.43 | 0.20 | 0.841 |  |
| +10 dB SNR |  |  |  |  | 0.42 |
|  | ENI index | 2.08 | 2.69 | 0.016 |  |
|  | NA ratio | 3.85 | 0.08 | 0.937 |  |
|  | NA speed | 0.511 | 1.59 | 0.131 |  |
|  | AR ratio | -33.87 | -0.75 | 0.463 |  |
|  | AR speed | -0.22 | -0.79 | 0.441 |  |
|  | AM ratio | 5.59 | 0.13 | 0.901 |  |
| +5 dB SNR |  |  |  |  | 0.51 |
|  | ENI index | 1.58 | 3.11 | 0.006 |  |
|  | NA ratio | -1.46 | -0.05 | 0.963 |  |
|  | NA speed | 0.36 | 1.69 | 0.109 |  |
|  | AR ratio | -0.86 | -0.03 | 0.977 |  |
|  | AR speed | -0.33 | -1.77 | 0.094 |  |
|  | AM ratio | -2.24 | -0.08 | 0.940 |  |

## REFERENCES

Adunka, O. F., Gantz, B. J., Dunn, C., Gurgel, R. K., & Buchman, C. A. (2018). Minimum reporting standards for adult cochlear implantation. Otolaryngology–Head and Neck Surgery, 159(2), 215–219. https://doi.org/10.1177/0194599818764329

Akaike, H. (1974). A new look at the statistical model identification. IEEE Transactions on Automatic Control, 19(6), 716–723. https://doi.org/10.1109/tac.1974.1100705

American Academy of Audiology. (2019). Clinical practice guidelines: Cochlear implants. https://www.audiology.org/wpcontent/uploads/2021/05/CochlearImplantPracticeGuidelines.pdf

Arjmandi, M. K., Jahn, K. N., & Arenberg, J. G. (2022). Single-Channel focused thresholds relate to vowel identification in pediatric and adult cochlear implant listeners. Trends in Hearing, 26, 233121652210953. https://doi.org/10.1177/23312165221095364

Berlin, C. I., Hood, L. J., Morlet, T., Wilensky, D., Li, L., Mattingly, K. R., Taylor-Jeanfreau, J., Keats, B. J. B., John, P. St., Montgomery, E., Shallop, J. K., Russell, B. A., & Frisch, S. A. (2010). Multi-site diagnosis and management of 260 patients with auditory neuropathy/dys-synchrony (auditory neuropathy spectrum disorder*). International Journal of Audiology, 49(1), 30–43. https://doi.org/10.3109/14992020903160892

Bernhard, N., Gauger, U., Romo Ventura, E., Uecker, F. C., Olze, H., Knopke, S., Hänsel, T., & Coordes, A. (2021). Duration of deafness impacts auditory performance after cochlear implantation: A meta-analysis. Laryngoscope Investigative Otolaryngology, 6(2), 291–301. https://doi.org/10.1002/lio2.528

Bierer, J. A. (2010). Probing the electrode-neuron interface with focused cochlear implant stimulation. Trends in Amplification, 14(2), 84–95. https://doi.org/10.1177/1084713810375249

Bittencourt, A. G., Torre, A. A. G. D., Bento, R. F., Tsuji, R. K., & Brito, R. de. (2012). Prelingual deafness: Benefits from cochlear implants versus conventional hearing aids. International Archives of Otorhinolaryngology, 16(3), 387–390. https://doi.org/10.7162/S1809-97772012000300014

Blamey, P., Arndt, P., Bergeron, F., Bredberg, G., Brimacombe, J., Facer, G., Larky, J., Lindström, B., Nedzelski, J., Peterson, A., Shipp, D., Staller, S., & Whitford, L. (1996). Factors affecting auditory performance of postlinguistically deaf adults using cochlear implants. Audiology & Neuro-Otology, 1(5), 293–306. https://doi.org/10.1159/000259212

Blamey, P., Artieres, F., Baskent, D., Bergeron, F., Beynon, A., Burke, E., Dillier, N., Dowell, R., Fraysse, B., Gallégo, S., Govaerts, P. J., Green, K., Huber, A. M., Kleine-Punte, A., Maat, B., Marx, M., Mawman, D., Mosnier, I., O?Connor, A. F., & O?Leary, S. (2013). Factors affecting auditory performance of postlinguistically deaf adults using cochlear implants: An update with 2251 patients. Audiology and Neurotology, 18(1), 36–47. https://doi.org/10.1159/000343189

Bo, D., Huang, Y., Wang, B., Lu, P., Chen, W., & Xu, Z. (2022). Auditory and speech outcomes of cochlear implantation in children with auditory neuropathy spectrum disorder: A systematic review and meta-analysis. Annals of Otology, Rhinology & Laryngology, 000348942210922. https://doi.org/10.1177/00034894221092201

Boisvert, I., Reis, M., Au, A., Cowan, R., & Dowell, R. C. (2020). Cochlear implantation outcomes in adults: A scoping review. PLOS ONE, 15(5), e0232421. https://doi.org/10.1371/journal.pone.0232421

Brown, C. J., Abbas, P. J., Etler, C. P., O’Brien, S., & Oleson, J. J. (2010). Effects of long-term use of a cochlear implant on the electrically evoked compound action potential. Journal of the American Academy of Audiology, 21(01), 005–015. https://doi.org/10.3766/jaaa.21.1.2

Buechner, A., Bardt, M., Haumann, S., Geissler, G., Salcher, R., & Lenarz, T. (2022). Clinical experiences with intraoperative electrocochleography in cochlear implant recipients and its potential to reduce insertion trauma and improve postoperative hearing preservation. PLOS ONE, 17(4), e0266077. https://doi.org/10.1371/journal.pone.0266077

Carlson, M. L., Sladen, D. P., Gurgel, R. K., Tombers, N. M., Lohse, C. M., & Driscoll, C. L. (2018). Survey of the american neurotology society on cochlear implantation: Part 1, candidacy assessment and expanding indications. Otology & Neurotology, 39(1), e12–e19. https://doi.org/10.1097/mao.0000000000001632

Delgutte, B. (1980). Representation of speech-like sounds in the discharge patterns of auditory-nerve fibers. The Journal of the Acoustical Society of America, 68(3), 843–857. https://doi.org/10.1121/1.384824

Delgutte, B. (1997). Auditory neural processing of speech in The Handbook of Phonetic Science. (W. J. Hardcastle & J. Laver, Eds.; pp. 507–538). Oxford: Blackwell.

Delgutte, B., & Kiang, N. Y. S. (1984). Speech coding in the auditory nerve: IV. Sounds with consonant-like dynamic characteristics. The Journal of the Acoustical Society of America, 75(3), 897–907. https://doi.org/10.1121/1.390599

Eisen, M. D., & Franck, K. H. (2004). Electrically evoked compound action potential amplitude growth functions and hiresolution programming levels in pediatric CII implant subjects. Ear and Hearing, 25(6), 528–538. https://doi.org/10.1097/00003446-200412000-00002

Finley, C. C., Holden, T. A., Holden, L. K., Whiting, B. R., Chole, R. A., Neely, G. J., Hullar, T. E., & Skinner, M. W. (2008). Role of electrode placement as a contributor to variability in cochlear implant outcomes. Otology & Neurotology, 29(7), 920–928. https://doi.org/10.1097/mao.0b013e318184f492

Garadat, S. N., Zwolan, T. A., & Pfingst, B. E. (2013). Using temporal modulation sensitivity to select stimulation sites for processor maps in cochlear implant listeners. Audiology and Neurotology, 18(4), 247–260. https://doi.org/10.1159/000351302

Gifford, R. H., Shallop, J. K., & Peterson, A. M. (2008). Speech recognition materials and ceiling effects: Considerations for cochlear implant programs. Audiology and Neurotology, 13(3), 193–205. https://doi.org/10.1159/000113510

Goudey, B., Plant, K., Kiral, I., Jimeno-Yepes, A., Swan, A., Gambhir, M., Büchner, A., Kludt, E., Eikelboom, R. H., Sucher, C., Gifford, R. H., Rottier, R., & Anjomshoa, H. (2021). A multicenter analysis of factors associated with hearing outcome for 2,735 adults with cochlear implants. Trends in Hearing, 25, 233121652110375. https://doi.org/10.1177/23312165211037525

Han, J. J., Suh, M.-W., Park, M. K., Koo, J.-W., Lee, J. H., & Oh, S. H. (2019). A predictive model for cochlear implant outcome in children with cochlear nerve deficiency. Scientific Reports, 9(1). https://doi.org/10.1038/s41598-018-37014-7

He, S., Abbas, P. J., Doyle, D. V., McFayden, T. C., & Mulherin, S. (2016). Temporal response properties of the auditory nerve in implanted children with auditory neuropathy spectrum disorder and implanted children with sensorineural hearing loss. Ear and Hearing, 37(4), 397–411. https://doi.org/10.1097/aud.0000000000000254

He, S., Shahsavarani, B. S., McFayden, T. C., Wang, H., Gill, K. E., Xu, L., Chao, X., Luo, J., Wang, R., & He, N. (2018). Responsiveness of the electrically stimulated cochlear nerve in children with cochlear nerve deficiency. Ear and Hearing, 39(2), 238–250. https://doi.org/10.1097/aud.0000000000000467

He, S., Skidmore, J., Conroy, S., Riggs, W. J., Carter, B. L., & Xie, R. (2022a). Neural adaptation of the electrically stimulated auditory nerve is not affected by advanced age in postlingually deafened, middle-aged, and elderly adult cochlear implant users. Ear & Hearing, 43(4), 1228–1244. https://doi.org/10.1097/aud.0000000000001184

He, S., Skidmore, J., & Carter, B. L. (2022b). Characteristics of the adaptation recovery function of the auditory nerve and its association with advanced age in postlingually deafened adult cochlear implant users. Ear & Hearing, Publish Ahead of Print. https://doi.org/10.1097/aud.0000000000001198

He, S., Skidmore, J., Carter, B. L., Lemeshow, S., & Sun, S. (2022c). Postlingually deafened adult cochlear implant users with prolonged recovery from neural adaptation at the level of the auditory nerve tend to have poorer speech perception performance. Ear & Hearing, Publish Ahead of Print. https://doi.org/10.1097/aud.0000000000001244

He, S., Skidmore, J., Chatterjee, M., Carter, B. L., & Yuan, Y. (2022d). Relationships between the auditory nerve sensitivity to amplitude modulation, perceptual amplitude modulation rate discrimination sensitivity and speech perception performance in postlingually deafened adult cochlear implant users. Ear and Hearing. [Conditionally Accepted].

He, S., Teagle, H. F. B., & Buchman, C. A. (2017). The electrically evoked compound action potential: From laboratory to clinic. Frontiers in Neuroscience, 11. https://doi.org/10.3389/fnins.2017.00339

Heutink, F., Verbist, B. M., van der Woude, W.-J., Meulman, T. J., Briaire, J. J., Frijns, J. H. M., Vart, P., Mylanus, E. A. M., & Huinck, W. J. (2021). Factors influencing speech perception in adults with a cochlear implant. Ear & Hearing, 42(4), 949–960. https://doi.org/10.1097/aud.0000000000000988

Hey, M., Neben, N., Stöver, T., Baumann, U., Mewes, A., Liebscher, T., Schüssler, M., Aschendorff, A., Wesarg, T., Büchner, A., Greenham, P., & Hoppe, U. (2020). Outcomes for a clinically representative cohort of hearing-impaired adults using the Nucleus® CI532 cochlear implant. European Archives of Oto-Rhino-Laryngology, 277(6), 1625–1635. https://doi.org/10.1007/s00405-020-05893-0

Holden, L. K., Finley, C. C., Firszt, J. B., Holden, T. A., Brenner, C., Potts, L. G., Gotter, B. D., Vanderhoof, S. S., Mispagel, K., Heydebrand, G., & Skinner, M. W. (2013). Factors affecting open-set word recognition in adults with cochlear implants. Ear & Hearing, 34(3), 342–360. https://doi.org/10.1097/aud.0b013e3182741aa7

Holden, L. K., Firszt, J. B., Reeder, R. M., Uchanski, R. M., Dwyer, N. Y., & Holden, T. A. (2016). Factors affecting outcomes in cochlear implant recipients implanted with a perimodiolar electrode array located in scala tympani. Otology & Neurotology, 37(10), 1662–1668. https://doi.org/10.1097/mao.0000000000001241

Hughes, M. L., Castioni, E. E., Goehring, J. L., & Baudhuin, J. L. (2012). Temporal response properties of the auditory nerve: Data from human cochlear-implant recipients. Hearing Research, 285(1-2), 46–57. https://doi.org/10.1016/j.heares.2012.01.010

James, C. J., Karoui, C., Laborde, M.-L., Lepage, B., Molinier, C.-É., Tartayre, M., Escudé, B., Deguine, O., Marx, M., & Fraysse, B. (2019). Early sentence recognition in adult cochlear implant users. Ear & Hearing, 40(4), 905–917. https://doi.org/10.1097/aud.0000000000000670

Johnson, D. H. (1980). The relationship between spike rate and synchrony in responses of auditory-nerve fibers to single tones. The Journal of the Acoustical Society of America, 68(4), 1115–1122. https://doi.org/10.1121/1.384982

Kaandorp, M. W., Smits, C., Merkus, P., Festen, J. M., & Goverts, S. T. (2017). Lexical-Access ability and cognitive predictors of speech recognition in noise in adult cochlear implant users. Trends in Hearing, 21, 233121651774388. https://doi.org/10.1177/2331216517743887

Kamakura, T., & Nadol, J. B. (2016). Correlation between word recognition score and intracochlear new bone and fibrous tissue after cochlear implantation in the human. Hearing Research, 339, 132–141. https://doi.org/10.1016/j.heares.2016.06.015

Kang, W. S., Lee, J. H., Lee, H. N., & Lee, K.-S. (2010). Cochlear implantations in young children with cochlear nerve deficiency diagnosed by MRI. Otolaryngology–Head and Neck Surgery, 143(1), 101–108. https://doi.org/10.1016/j.otohns.2010.03.016

Kraaijenga, V. J. C., Derksen, T. C., Stegeman, I., & Smit, A. L. (2018). The effect of side of implantation on unilateral cochlear implant performance in patients with prelingual and postlingual sensorineural hearing loss: A systematic review. Clinical Otolaryngology: Official Journal of ENT-UK ; Official Journal of Netherlands Society for Oto-Rhino-Laryngology & Cervico-Facial Surgery, 43(2), 440–449. https://doi.org/10.1111/coa.12988

Kraus, N., Bradlow, A. R., Cheatham, M. A., Cunningham, J., King, C. D., Koch, D. B., Nicol, T. G., McGee, T. J., Stein, L. K., & Wright, B. A. (2000). Consequences of neural asynchrony: A case of auditory neuropathy. Journal of the Association for Research in Otolaryngology, 1(1), 33–45. https://doi.org/10.1007/s101620010004

Lenarz, M., Sönmez, H., Joseph, G., Büchner, A., & Lenarz, T. (2012). Cochlear implant performance in geriatric patients. The Laryngoscope, 122(6), 1361–1365. https://doi.org/10.1002/lary.23232

Liang, C., Wenstrup, L. H., Samy, R. N., Xiang, J., & Zhang, F. (2020). The effect of side of implantation on the cortical processing of frequency changes in adult cochlear implant users. Frontiers in Neuroscience, 14. https://doi.org/10.3389/fnins.2020.00368

Long, C. J., Holden, T. A., McClelland, G. H., Parkinson, W. S., Shelton, C., Kelsall, D. C., & Smith, Z. M. (2014). Examining the electro-neural interface of cochlear implant users using psychophysics, CT scans, and speech understanding. Journal of the Association for Research in Otolaryngology, 15(2), 293–304. https://doi.org/10.1007/s10162-013-0437-5

Mussoi, B. S. S., & Brown, C. J. (2019). Age-Related changes in temporal resolution revisited. Ear and Hearing, 40(6), 1328–1344. https://doi.org/10.1097/aud.0000000000000732

Myers, K., & Nicholson, N. (2021). Cochlear implant behavioral outcomes for children with auditory neuropathy spectrum disorder: A mini-systematic review. American Journal of Audiology, 30(3), 777–789. https://doi.org/10.1044/2021_aja-20-00175

National Institute on Deafness and Other Communication Disorders. (2021). Cochlear Implants. (NIH Publication No. 00-4798). Washington, DC: U.S. Government Printing Office.

Peterson, G. E., & Lehiste, I. (1962). Revised CNC lists for auditory tests. Journal of Speech and Hearing Disorders, 27(1), 62–70. https://doi.org/10.1044/jshd.2701.62

Pfingst, B. E., Colesa, D. J., Swiderski, D. L., Hughes, A. P., Strahl, S. B., Sinan, M., & Raphael, Y. (2017). Neurotrophin gene therapy in deafened ears with cochlear implants: Long-term effects on nerve survival and functional measures. Journal of the Association for Research in Otolaryngology, 18(6), 731–750. https://doi.org/10.1007/s10162-017-0633-9

Pfingst, B. E., Zhou, N., Colesa, D. J., Watts, M. M., Strahl, S. B., Garadat, S. N., Schvartz-Leyzac, K. C., Budenz, C. L., Raphael, Y., & Zwolan, T. A. (2015). Importance of cochlear health for implant function. Hearing Research, 322, 77–88. https://doi.org/10.1016/j.heares.2014.09.009

R Core Team. (2022). R: A language and environment for statistical computing. R Foundation for Statistical Computing; Vienna, Austria. https://www.R-project.org

Ramekers, D., Versnel, H., Strahl, S. B., Klis, S. F. L., & Grolman, W. (2015). Recovery characteristics of the electrically stimulated auditory nerve in deafened guinea pigs: Relation to neuronal status. Hearing Research, 321, 12–24. https://doi.org/10.1016/j.heares.2015.01.001

Ramekers, D., Versnel, H., Strahl, S. B., Smeets, E. M., Klis, S. F. L., & Grolman, W. (2014). Auditory-Nerve responses to varied inter-phase gap and phase duration of the electric pulse stimulus as predictors for neuronal degeneration. Journal of the Association for Research in Otolaryngology, 15(2), 187–202. https://doi.org/10.1007/s10162-013-0440-x

Rance, G., Barker, E., Mok, M., Dowell, R., Rincon, A., & Garratt, R. (2007). Speech perception in noise for children with auditory neuropathy/dys-synchrony type hearing loss. Ear and Hearing, 28(3), 351–360. https://doi.org/10.1097/aud.0b013e3180479404

Rasmussen, K. M. B., West, N. C., Bille, M., Sandvej, M. G., & Cayé-Thomasen, P. (2022). Cochlear implantation improves both speech perception and patient-reported outcomes: A prospective follow-up study of treatment benefits among adult cochlear implant recipients. Journal of Clinical Medicine, 11(8), 2257. https://doi.org/10.3390/jcm11082257

Riggs, W. J., Vaughan, C., Skidmore, J., Conroy, S., Pellittieri, A., Carter, B. L., Stegman, C. J., & He, S. (2021). The sensitivity of the electrically stimulated auditory nerve to amplitude modulation cues declines with advanced age. Ear & Hearing, 42(5). https://doi.org/10.1097/aud.0000000000001035

Schvartz-Leyzac, K. C., Holden, T. A., Zwolan, T. A., Arts, H. A., Firszt, J. B., Buswinka, C. J., & Pfingst, B. E. (2020). Effects of electrode location on estimates of neural health in humans with cochlear implants. Journal of the Association for Research in Otolaryngology, 21(3), 259–275. https://doi.org/10.1007/s10162-020-00749-0

Schvartz-Leyzac, K. C., & Pfingst, B. E. (2016). Across-site patterns of electrically evoked compound action potential amplitude-growth functions in multichannel cochlear implant recipients and the effects of the interphase gap. Hearing Research, 341, 50–65. https://doi.org/10.1016/j.heares.2016.08.002

Seyyedi, M., & Nadol Jr, J. B. (2014). Intracochlear inflammatory response to cochlear implant electrodes in humans. Otology & Neurotology, 35(9), 1545–1551. https://doi.org/10.1097/mao.0000000000000540

Shallop, J. K. (2002). Auditory neuropathy/dys-synchrony in adults and children. Seminars in Hearing, 23(3), 215–224. https://doi.org/10.1055/s-2002-34474

Shepherd, R. K., Roberts, L. A., & Paolini, A. G. (2004). Long-term sensorineural hearing loss induces functional changes in the rat auditory nerve. European Journal of Neuroscience, 20(11), 3131–3140. https://doi.org/10.1111/j.1460-9568.2004.03809.x

Skidmore, J., Carter, B. L., Riggs, W. J., & He, S. (2022a). The effect of advanced age on the electrode-neuron interface in cochlear implant users. Ear & Hearing, 43(4), 1300–1315. https://doi.org/10.1097/aud.0000000000001185

Skidmore, J., Ramekers, D., Colesa, D. J., Schvartz-Leyzac, K. C., Pfingst, B. E., & He, S. (2022b). A broadly applicable method for characterizing the slope of the electrically evoked compound action potential amplitude growth function. Ear & Hearing, 43(1), 150–164. https://doi.org/10.1097/aud.0000000000001084

Skidmore, J., Xu, L., Chao, X., Riggs, W. J., Pellittieri, A., Vaughan, C., Ning, X., Wang, R., Luo, J., & He, S. (2021). Prediction of the functional status of the cochlear nerve in individual cochlear implant users using machine learning and electrophysiological measures. Ear & Hearing, 42(1), 180–192. https://doi.org/10.1097/aud.0000000000000916

Spahr, A. J., Dorman, M. F., Litvak, L. M., Van Wie, S., Gifford, R. H., Loizou, P. C., Loiselle, L. M., Oakes, T., & Cook, S. (2012). Development and validation of the AzBio sentence lists. Ear & Hearing, 33(1), 112–117. https://doi.org/10.1097/aud.0b013e31822c2549

Starr, A., Sininger, Y., Winter, M., Derebery, M. J., Oba, S., & Michalewski, H. J. (1998). Transient deafness due to temperature-sensitive auditory neuropathy. Ear and Hearing, 19(3), 169–179. https://doi.org/10.1097/00003446-199806000-00001

Tejani, V. D., Abbas, P. J., & Brown, C. J. (2017). Relationship between peripheral and psychophysical measures of amplitude modulation detection in cochlear implant users. Ear and Hearing, 38(5), e268–e284. https://doi.org/10.1097/aud.0000000000000417

van Eijl, R. H. M., Buitenhuis, P. J., Stegeman, I., Klis, S. F. L., & Grolman, W. (2016). Systematic review of compound action potentials as predictors for cochlear implant performance. The Laryngoscope, 127(2), 476–487. https://doi.org/10.1002/lary.26154

Walker, E., McCreery, R., Spratford, M., & Roush, P. (2016). Children with auditory neuropathy spectrum disorder fitted with hearing aids applying the american academy of audiology pediatric amplification guideline: Current practice and outcomes. Journal of the American Academy of Audiology, 27(3), 204–218. https://doi.org/10.3766/jaaa.15050

Wilson, B. S., Finley, C. C., Lawson, D. T., Wolford, R. D., Eddington, D. K., & Rabinowitz, W. M. (1991). Better speech recognition with cochlear implants. Nature, 352(6332), 236–238. https://doi.org/10.1038/352236a0

Wilson, B. S., Finley, C. C., Lawson, D. T., & Zerbi, M. (1997). Temporal representations with cochlear implants. The American Journal of Otology, 18(6), S30–34.

Zeng, F.-G., & Liu, S. (2006). Speech perception in individuals with auditory neuropathy. Journal of Speech, Language, and Hearing Research, 49(2), 367–380. https://doi.org/10.1044/1092-4388(2006/029)

Zhang, F., Benson, C., Murphy, D., Boian, M., Scott, M., Keith, R., Xiang, J., & Abbas, P. (2013). Neural adaptation and behavioral measures of temporal processing and speech perception in cochlear implant recipients. PLoS ONE, 8(12), e84631. https://doi.org/10.1371/journal.pone.0084631

Zhao, E. E., Dornhoffer, J. R., Loftus, C., Nguyen, S. A., Meyer, T. A., Dubno, J. R., & McRackan, T. R. (2020). Association of patient-related factors with adult cochlear implant speech recognition outcomes: A meta-analysis. JAMA Otolaryngology–Head & Neck Surgery, 146(7), 613–620. https://doi.org/10.1001/jamaoto.2020.0662

